# Trunk control in and out of an episode of recurrent low back pain during the Balance-Dexterity Task

**DOI:** 10.1101/2022.09.23.22280309

**Authors:** Hai-Jung Steffi Shih, Joyce Ai, Justin Abe, Jiaxi Tang, K. Michael Rowley, Linda R. Van Dillen, Kornelia Kulig

## Abstract

We investigated motor control strategies utilized by individuals with recurrent low back pain (rLBP) during active pain and remission periods as well as by back-healthy controls using the Balance-Dexterity Task. Nineteen young adults with rLBP were tested first when they were in pain and then again in symptom remission, and 19 matched controls were also tested. Trunk kinematic coupling and muscle co-activation were examined while participants performed the task by standing on one leg while compressing a spring with a maximum consistent force with the other leg. We found a decreased bilateral external oblique co-activation and a further reduced trunk coupling during the spring condition of the task compared to in a stable block condition in people with rLBP compared to back-healthy individuals. When individuals were in active pain, they exhibited more co-activation than when they were in remission, but the co-activation was not greater than in back-healthy individuals.

## 1. Introduction

Over eighty percent of adults experience low back pain (LBP) at some point in their lives and the prevalence of LBP is increasing as the population ages.^1^ High disability^2^ associated with LBP is largely attributed to its chronic and recurrent nature.^3^ Development of clinical intervention and prevention strategies is challenging because the majority of LBP is non-specific - without identifiable pathology.^4^ A systematic review reported that the only significant predictor of LBP recurrence is a history of previous episodes,^5^ however, studies included did not examine motor control related factors. Aberrant motor control, including muscle activation and trunk kinematics, is frequently reported in individuals with LBP, which is not surprising given LBP is often influenced by movement.^6^

The abundant literature on motor control in patients with LBP produced variable task- and population-dependent results. To address the divergence in findings, a recent commentary proposed that persons with LBP differ in motor control compared to back-healthy individuals, and the difference could be seen at two ends of the spectrum: “tight” control and “loose” control strategies.^7^ “Tight” control of the trunk constrains movement to avoid pain or injury, and is indicated by increased muscle co-activation and trunk stiffness; “Loose” control of the trunk decreases muscle activation to prevent pain provocation and reduce tissue loading, and is indicated by reduced muscle co-activation and trunk stiffness.^7^ Both strategies may have consequences that could contribute to further pain - “tight” control may lead to reduced movement variability and increased compression forces, resulting in greater, repetitive loading on spinal tissues; “loose” control may lead to excessive shearing forces between spinal segments, resulting in tissue strains.^7^

The “tight” and “loose” motor control strategies were speculated to be demonstrated in different tasks.^8^ For example, a more threatening task such as sudden perturbation to the trunk had elicited increased trunk stiffness in people with recurrent LBP.^9^ On the other hand, a less threatening task such as self-initiated forward bend to pick up an object had resulted in more lumbar motion in the early part of the task in people with chronic LBP compared to back-healthy people.^10^ As many studies on motor control in people with LBP used discrete perturbation tasks,^9,11^ more research using continuous, low-threat tasks are warranted to further understand task-dependent motor control strategies.

The Balance-Dexterity Task (BDT)^12^ combines a single-leg balance task with the Lower-Extremity Dexterity Test,^13^ and serves as a self-initiated, continuous, and less-threatening task to probe trunk control strategies in people with LBP. Previous study using the BDT by Rowley et al. demonstrated reduced trunk coupling (more dissociated thorax and pelvis motion) in people in remission from recurrent LBP compared to back-healthy controls.^14^ This “loose” control strategy elicited by the less-threatening task concurs with previous literature supporting task-dependent divergence of motor control strategies in people with LBP. However, it is unknown how pain status may have affected trunk control strategies, as it is well-established that people move differently in pain.^15^ While there is a handful of studies examining people with a history of LBP without current symptoms,^14,16^ a direct comparison in and out of pain within the same cohort is warranted to further understand how motor control strategies are influenced by pain status.

The purpose of this study was to compare motor control strategies in people with recurrent LBP in and out of a painful episode, and back-healthy young adults using the BDT. We hypothesized that people with recurrent LBP would exhibit reduced trunk kinematic coupling during symptom remission compared to during a painful episode, as well as compared to back-healthy controls. We also hypothesized that people with recurrent LBP would exhibit reduced trunk muscle co-activation during symptom remission compared to during a painful episode, as well as compared to back-healthy controls.

## 2. Methods

### 2.1 Participants

Nineteen participants with recurrent low back pain (rLBP) along with 19 matched back-healthy individuals (CTRL) were recruited with approval by the Institutional Review Board of the University of Southern California (Table 1). All participants provided written informed consent after passing inclusion and exclusion criteria (Table 2).

**Table 1.**
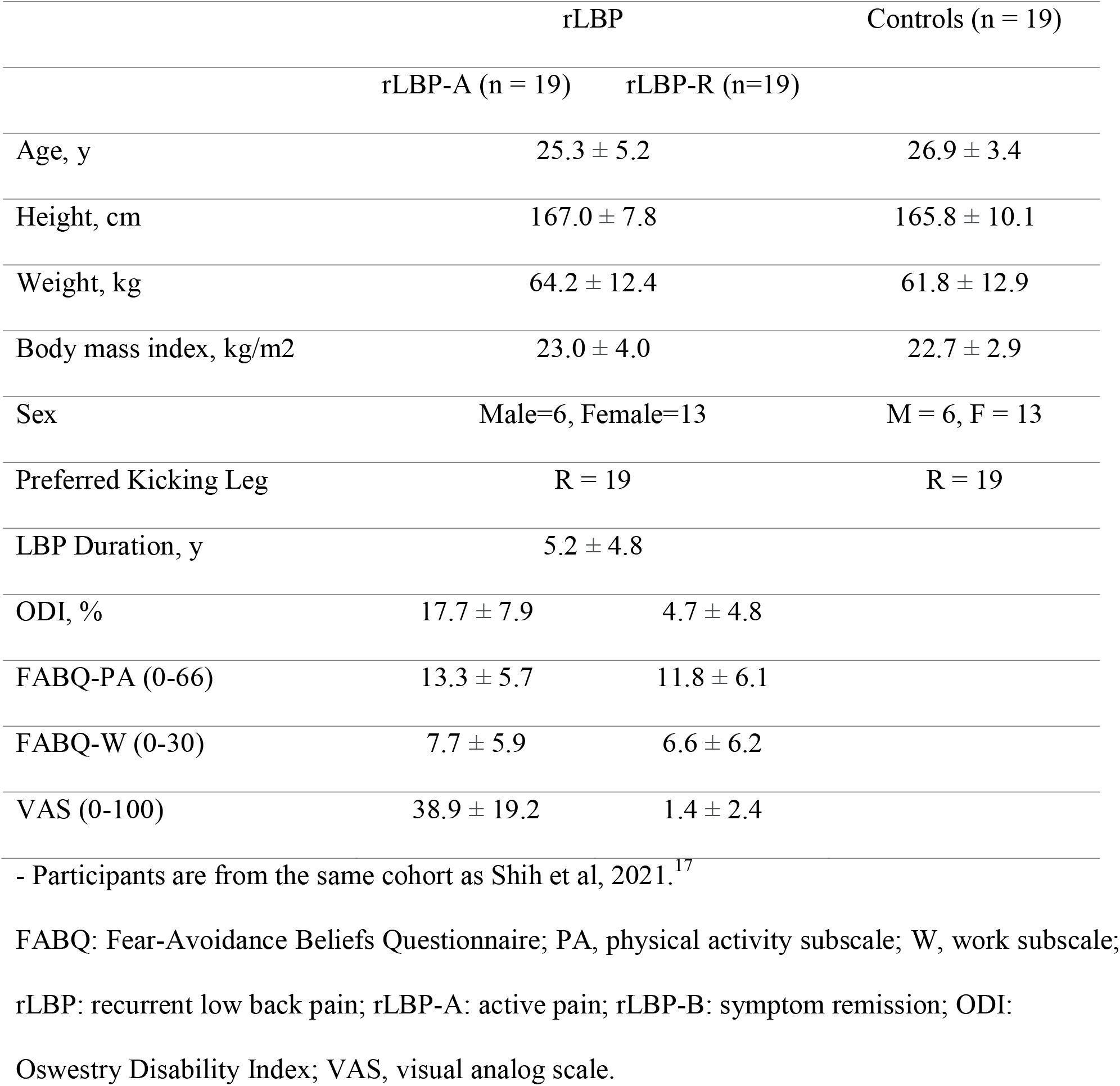
Participant characteristics (mean ± SD)

**Table 2.**
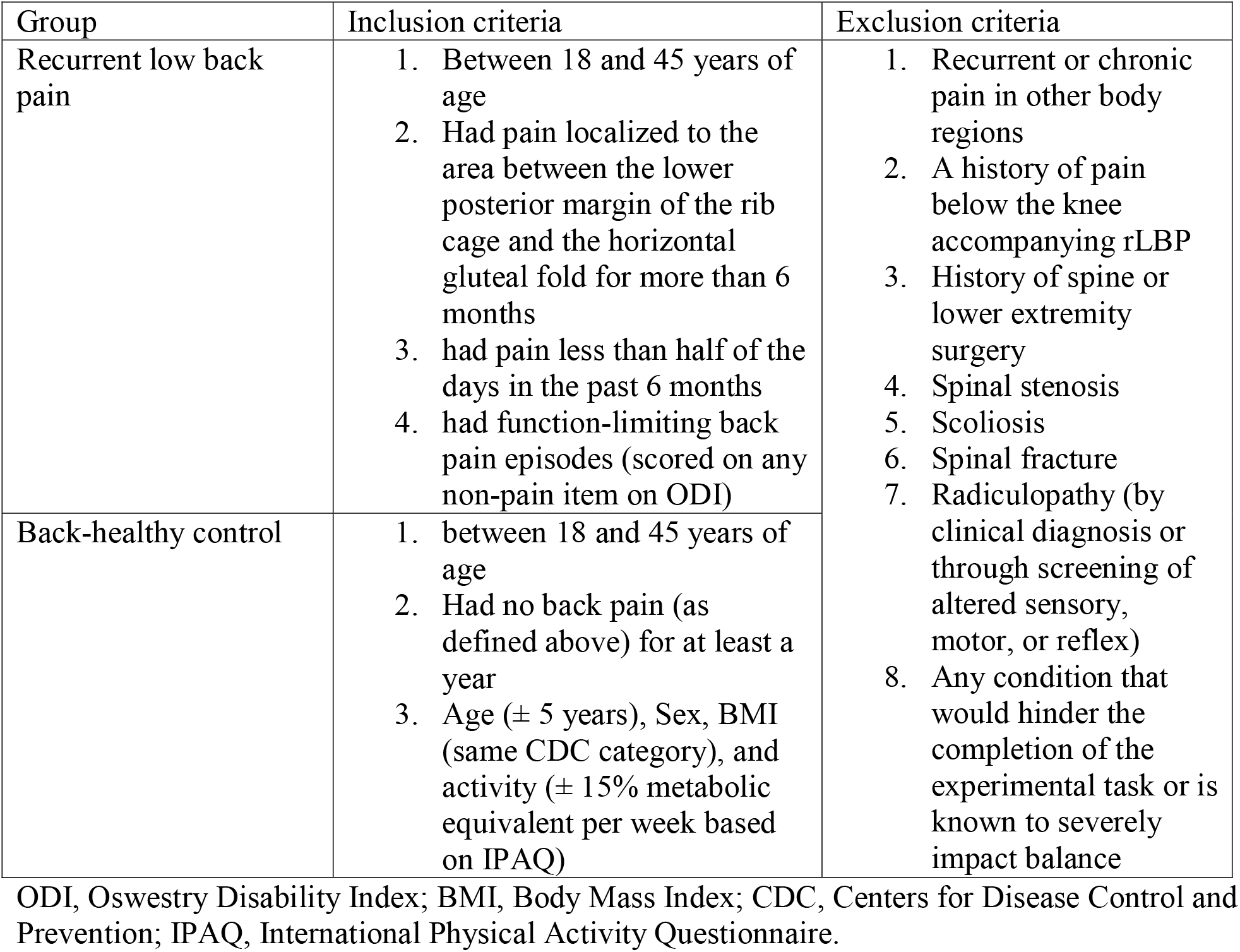
Inclusion and exclusion criteria.

Individuals with recurrent LBP were tested first in pain, then out of pain. They were considered to be in pain when the pain had persisted for more than 24 hours, with a score of ≥ 2/10 on the numeric rating scale.^18^ They were considered to be out of pain when their pain was <1/10 on numeric rating scale for more than 24 hours. Due to high test-retest reliability in our pilot study,^19^ back-healthy individuals were only tested once.

### 2.2 Instrumentation

Participants were instrumented with a lower extremity and trunk retroreflective marker set^19^ and surface electromyography (EMG) on 4 muscle groups bilaterally: rectus abdominis, external oblique, longissimus, and gluteus medius. Kinematic data were captured by an 11-camera Oqus System at 125 Hz (Qualisys; Gothenburg, Sweden); force data were captured by two force plates at 1500 Hz (Advanced Medical Technology Inc.; Watertown, MA); EMG data were captured by a wireless EMG system (Noraxon U.S.A, Inc.; Scottsdale, AZ) at 1500 Hz.

A custom-made device was used to introduce the BDT, consisting of a spring (Compression Spring Model 805; Century Spring Corporation, Commerce, CA) mounted between two 3 mm boards, with the bottom board attached to a force plate on the ground (Fig 1).^12^ The spring can be replaced by a rigid block of the same height to offer a stable condition for comparison. The BDT is an adaptation of the Lower-Extremity Dexterity Task^13^ by removing the pelvis and forearm support to include challenges to balance and trunk control.

**Fig 1.**
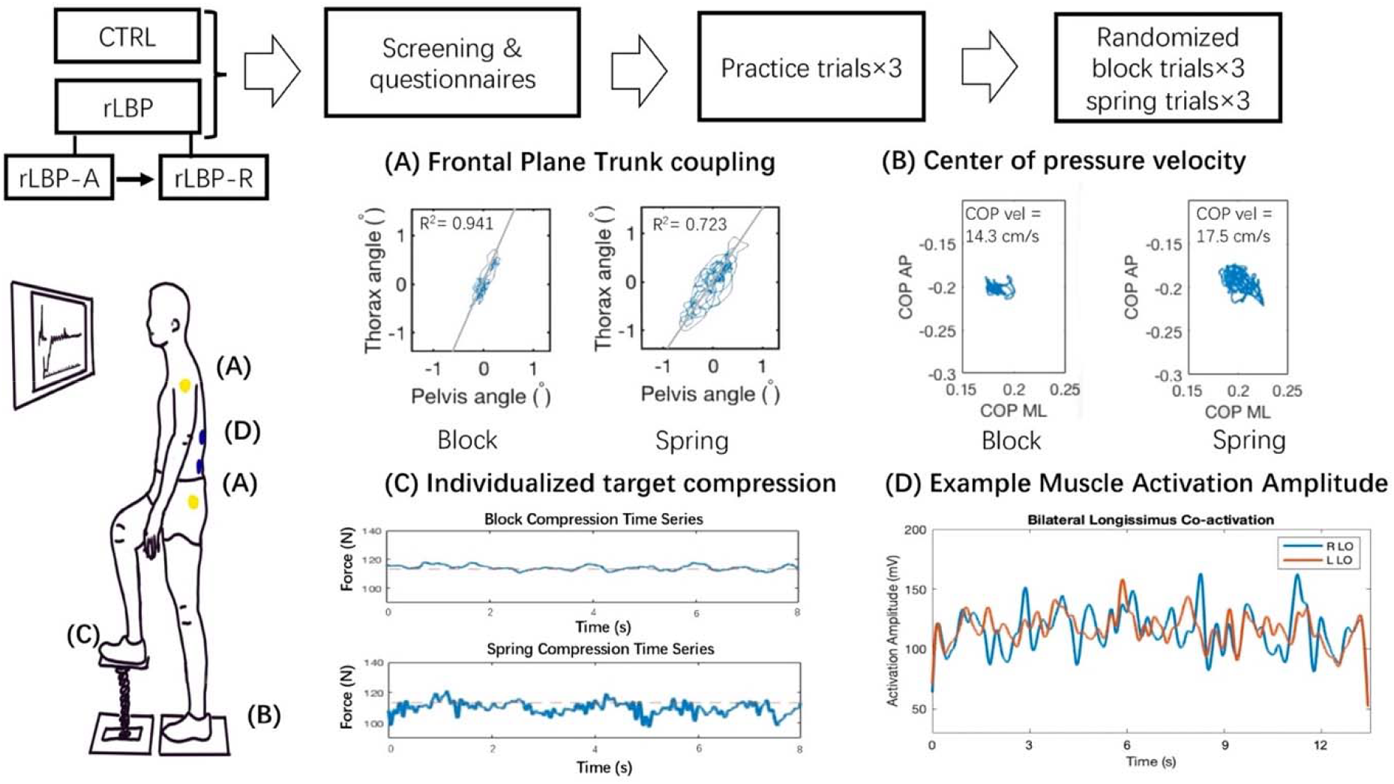
Overview of experimental procedures and the Balance-Dexterity task. Figure-top: flow chart of procedures in the laboratory. Figure-left: Balance-Dexterity task set-up, participant’s dexterity limb (preferred kicking leg) and supporting limb are placed anterior-posteriorly on individual force plates. Vertical ground reaction force collected under the dexterous limb (transmitted by either a spring or a stable block) is displayed relative to an individualized target compression force on the screen to provide visual feedback. Data examples: (A) Frontal plane trunk coupling R^2^ derived from thorax and pelvis segment angles, (B) Center of pressure velocity collected under the supporting limb, (C) Compression force outcome measure around individualized target under the dexterity limb, and (D) Muscle activation amplitude of two trunk muscles. CTRL, control; rLBP-A, recurrent low back pain – active pain; rLBP-R, recurrent low back pain – in remission; COP, center of pressure; AP, anterior-posterior; ML, medial-lateral.

### 2.3 Experimental Procedures

Prior to the experimental task, all participants completed the International Physical Activity Questionnaire (IPAQ),^20^ and those with rLBP completed the Oswestry Disability Index (ODI),^21^ Fear Avoidance Beliefs Questionnaire (FABQ),^22^ and Visual Analog Scale (VAS) for pain level at the time of data collection.

Participants were then introduced to the BDT (Fig 1). Participants placed their preferred kicking foot on a spring and compressed downwards for 30 seconds while observing real-time feedback of the vertical compression force under the spring on a screen in front of them. They were instructed to do the following: “While standing on one leg, compress the spring so that the line is first as high, then as stable as possible”. Participants completed 3 practice trials. Individualized target compression forces were calculated by taking the average force of the best performed trial (i.e. stable and highest). Participants then completed 3 ‘spring trials’ and 3 trials with a stable block in place of the spring (‘block trials’) in a randomized order. Their target compression force along with real-time visual feedback of vertical ground reaction force was shown on a screen before them, with the following instructions: “While standing on one leg, compress this spring/block so that the moving line (feedback) is as stable as possible directly over the dotted line (target).”

### 2.4 Signal Processing

All trials were truncated down to the middle 15 seconds to avoid the effects of initiation and termination of the task. Kinematic data were low-pass filtered at 10 Hz and force data at 50 Hz using a dual-pass 4^th^-order Butterworth filter in Visual 3D (C-motion, Inc, Germantown, MD). All subsequent data processing was conducted in Matlab (The Mathworks, Inc., Natick, MA).

#### 2.4.1 Task performance (dexterity and balance control)

Task performance was indexed by processed data extracted from the force plate. To describe dexterous control, root-mean-squared-error (RMSE) of the force in the vertical direction from the spring was used as an indicator. The RMSE was calculated relative to the individualized target compression force. A high RMSE would indicate poor dexterous control of the limb controlling compression force while a low RMSE would indicate good dexterous control. Average center of pressure (COP) resultant velocity from the support limb was used to describe balance control.^23^ A high COP velocity would indicate more postural sway, and a low COP velocity would indicate less postural sway and generally associated with better balance control.

#### 2.4.2 Frontal Plane Trunk coupling

A coefficient of determination (R^2^) was calculated between the thorax and pelvis segment angles in the frontal plane on an angle-angle plot as an indicator for trunk coupling. Based on Rowley et al, this metric provides similar information to a vector-coding analysis.^12^ A high R^2^ would indicate that thorax and pelvis are more likely to move in the same direction and a low R^2^ would indicate that there is more independent motion of the two segments.

#### 2.4.3 Muscle activation (amplitude and exploratory co-activation)

EMG data were 1) band-pass filtered between 30-500 Hz for longissimus, rectus abdominis, and external oblique to eliminate heart rate contamination,^24^ and between 20-500Hz for gluteus medius, using a dual-pass 4th order Butterworth filter, 2) full wave rectified with a 500ms moving window, and 3) normalized to the muscle’s average activation during the stable block trials to allow for comparisons. Referencing submaximal muscle contractions have been found to be more reliable in people with LBP than referencing maximal muscle contractions.^25^ Activation amplitude for each muscle was calculated by averaging the signal across the truncated 15-seconds trial duration and across the three individual trials for block and spring conditions.

Muscle co-activation was explored for the following 6 muscle pairings, which were selected to cover co-activation across the frontal, sagittal, and transverse planes of the trunk and the frontal plane of the pelvis:

- Bilateral Longissimus (LO)
- Bilateral Rectus Abdominis (RA)
- Bilateral External Oblique (EO)
- Bilateral Gluteus Medius (GM)
- Right LO- Right RA
- Left LO- Left RA

The co-activation index was calculated using the following equation^26^ that has been shown to have good sensitivity^27^:

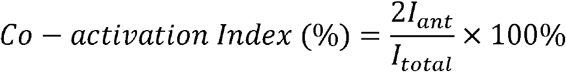

Where *I*_ant_ represents the area of total antagonistic activity,

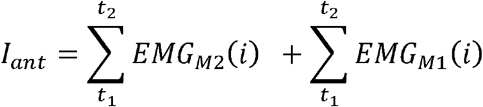

And *I*_*Total*_ represents the sum of EMG activity of the muscle pairing:

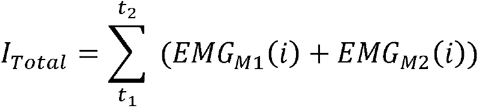

### 2.5 Statistical Analysis

R was used for all statistical analyses.^28^ Data were first plotted to visually inspect for normality and outliers. Descriptive statistics were performed, and Welch’s t-tests were conducted to ensure comparable participant characteristics. The analysis was performed in two steps. First, vertical force RMSE, COP average velocity, trunk coupling (R^2^) and muscle co-activation indices were compared within the rLBP group, between active pain (rLBP-A) and symptom remission (rLBP-R) conditions. A linear mixed-effects model with random intercept and slope was used to investigate the effect of pain status (rLBP-A vs. rLBP-R) and condition (block vs. spring), along with the interaction between pain status and condition. Individual participants were treated as random effects. Second, if there was a significant main effect of pain status or an interaction effect, rLBP-A and rLBP-R data were compared to the back-healthy control group (CTRL) separately. If these effects were non-significant, data for rLBP-A and rLBP-R were pooled (rLBP-Pooled) and compared to CTRL. The same 2-step procedure was carried out for activation amplitude of each muscle, except only the spring condition was included since these measures were normalized to the block condition. The level of significance was set at ✉ = 0.05.

## 3. Results

### 3.1 BDT Performance

#### 3.1.1 Dexterity control

No difference was found in target compression force between groups (p=.863). There were no differences in RMSE between rLBP-A and r-LBP-R (p=.363), so the rLBP data were pooled and compared with CTRL, and there was no difference between these groups (p=.421) (Fig 2(b)). RMSE in the spring condition was significantly greater than the block condition across groups (p<.001), indicating poorer task performance.

**Fig 2:**
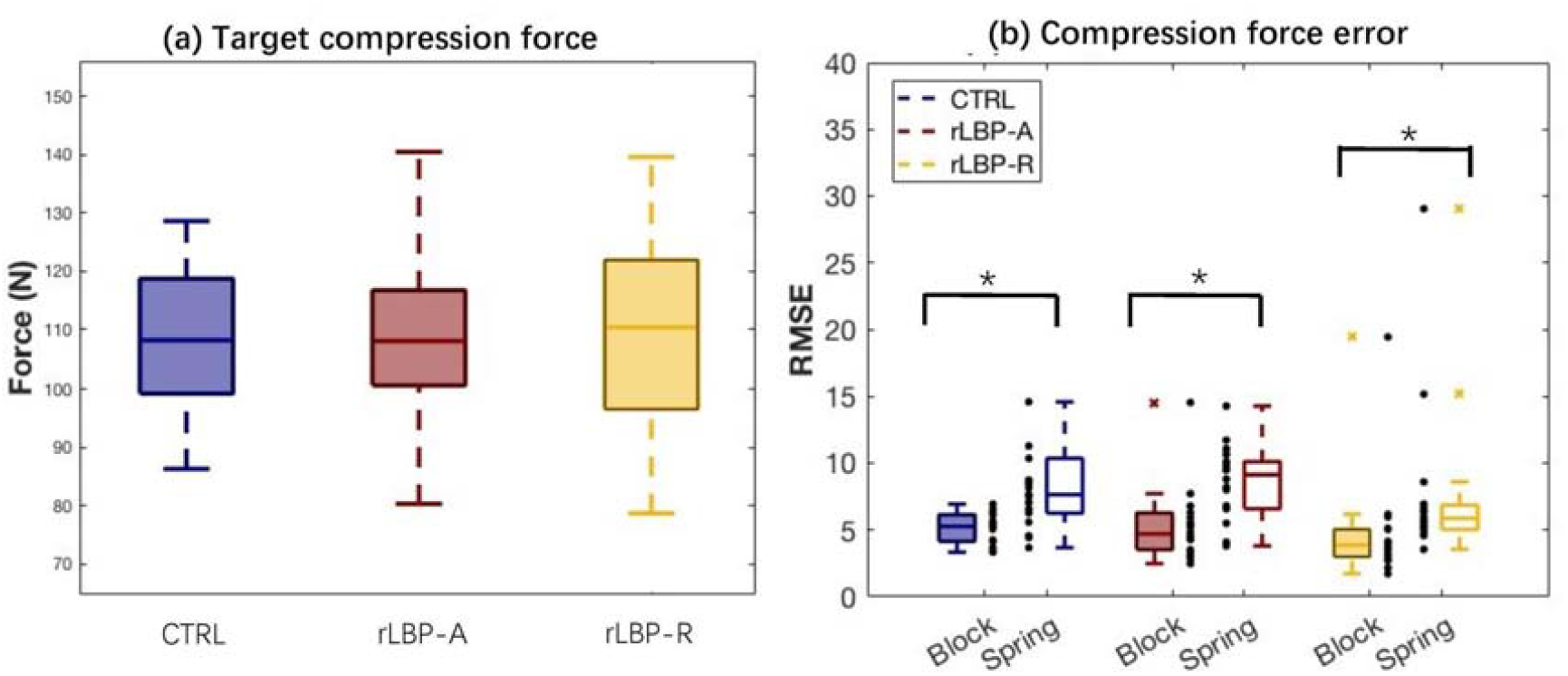
(a)Target compression force was calculated by the greatest and most stable force trajectory of the three practice trials. (b)Compression force error was calculated as root mean square error relative to target compression force. Individual data points are presented as dots alongside box plots representing group data. One outlier with high error in the CTRL group was removed visually (but not statistically) to retain the clarity of the figure. CTRL, control; rLBP-A, recurrent low back pain in active pain; rLBP-R, recurrent low back pain in remission; RMSE, root mean square error.

#### 3.1.2 Balance control

rLBP-R had reduced COP velocity compared to rLBP-A (p=.029), as well as compared to CTRL (Group main effect p=.011) (Fig 4). COP velocity in the spring condition was significantly greater than the block condition across all groups (Condition main effect p<.001).

### 3.2 Frontal Plane Trunk Coupling

There were no differences in trunk coupling between rLBP-A and rLBP-R (p=.695) or between rLBP-pooled and CTRL (p=.327) (Fig 3). Reduced trunk coupling, indicating more dissociated trunk and pelvis motion, in the spring condition compared to the block condition was identified in the ANOVA comparing rLBP-A and rLBP-R (Condition main effect p=.018). Due to a non-significant group main effect between rLBP-A and rLBP-R, we pooled rLBP-R and rLBP-A and performed an ANOVA for rLBP-pooled and CTRL and did not find a condition main effect or interaction effect.

**Fig 3:**
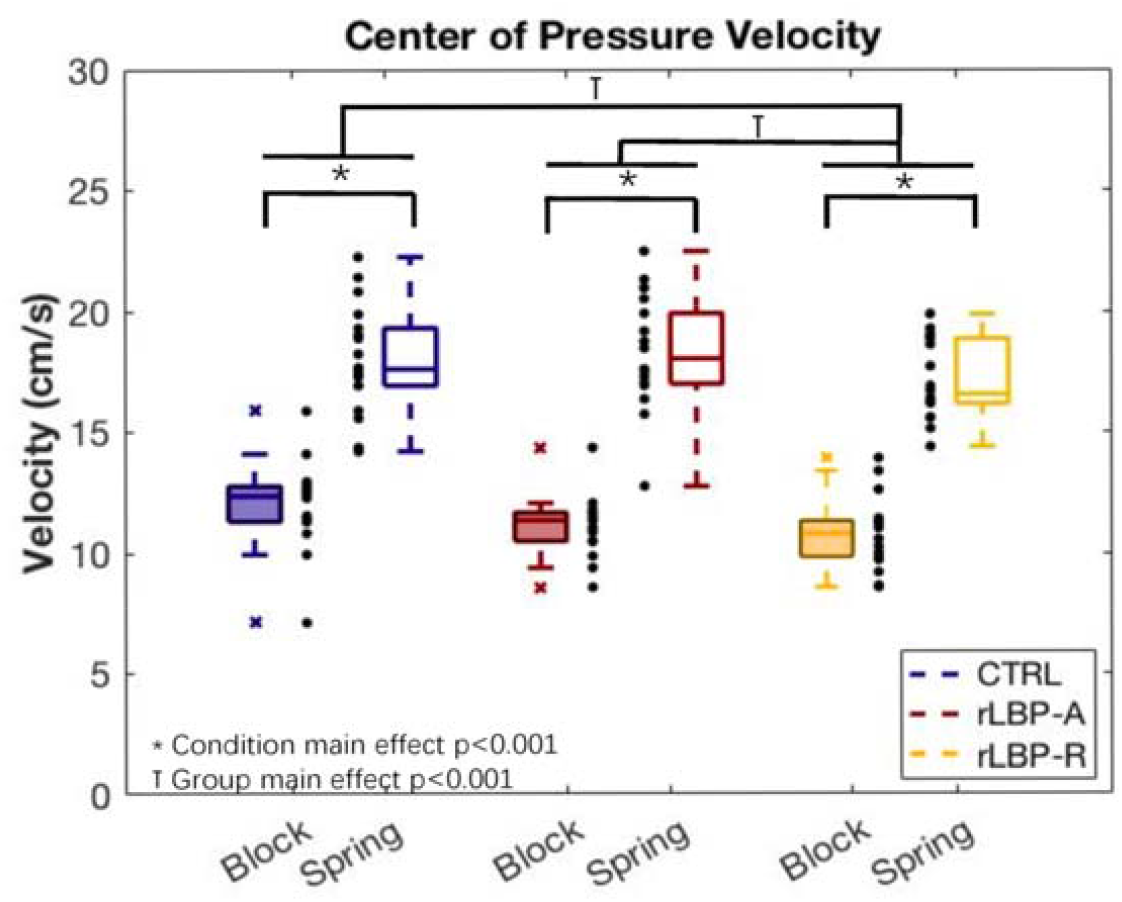
Center of pressure velocity. COP, center of pressure; CTRL, control; rLBP-A, recurrent low back pain in active pain; rLBP-R, recurrent low back pain in remission

### 3.3 Muscle Activation Amplitude

Comparisons across all muscles in spring condition trials revealed no differences between rLBP-A and rLBP-R (p=.225-.961), as well as between rLBP-Pooled and CTRL (p=.353-.980) (Fig 5). The activation amplitudes for all muscles in the spring condition trials were significantly greater than the block condition trials (p<.001-.031) across all groups.

**Fig 4:**
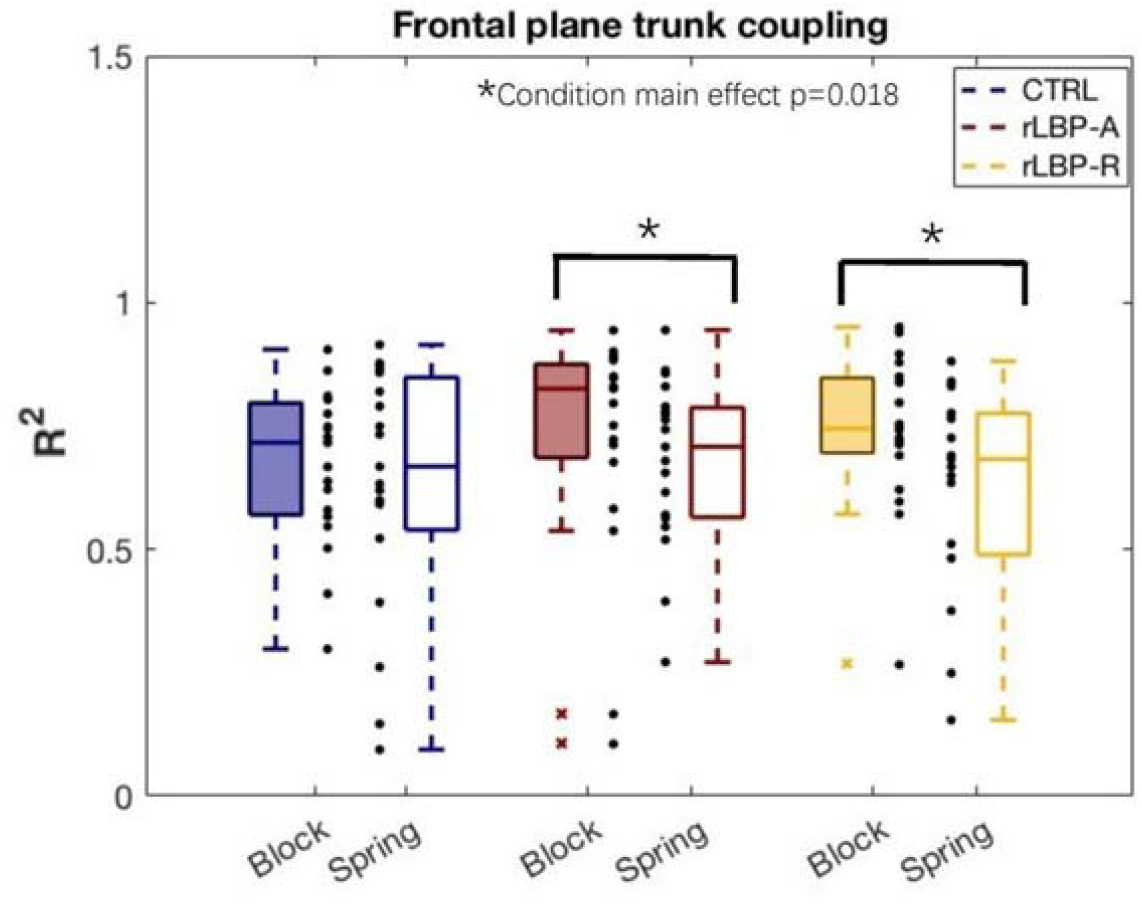
Frontal plane trunk coupling as indicated by coefficient of determination (R^2^). CTRL, control; rLBP-A, recurrent low back pain in active pain; rLBP-R, recurrent low back pain in remission.

**Fig 5:**
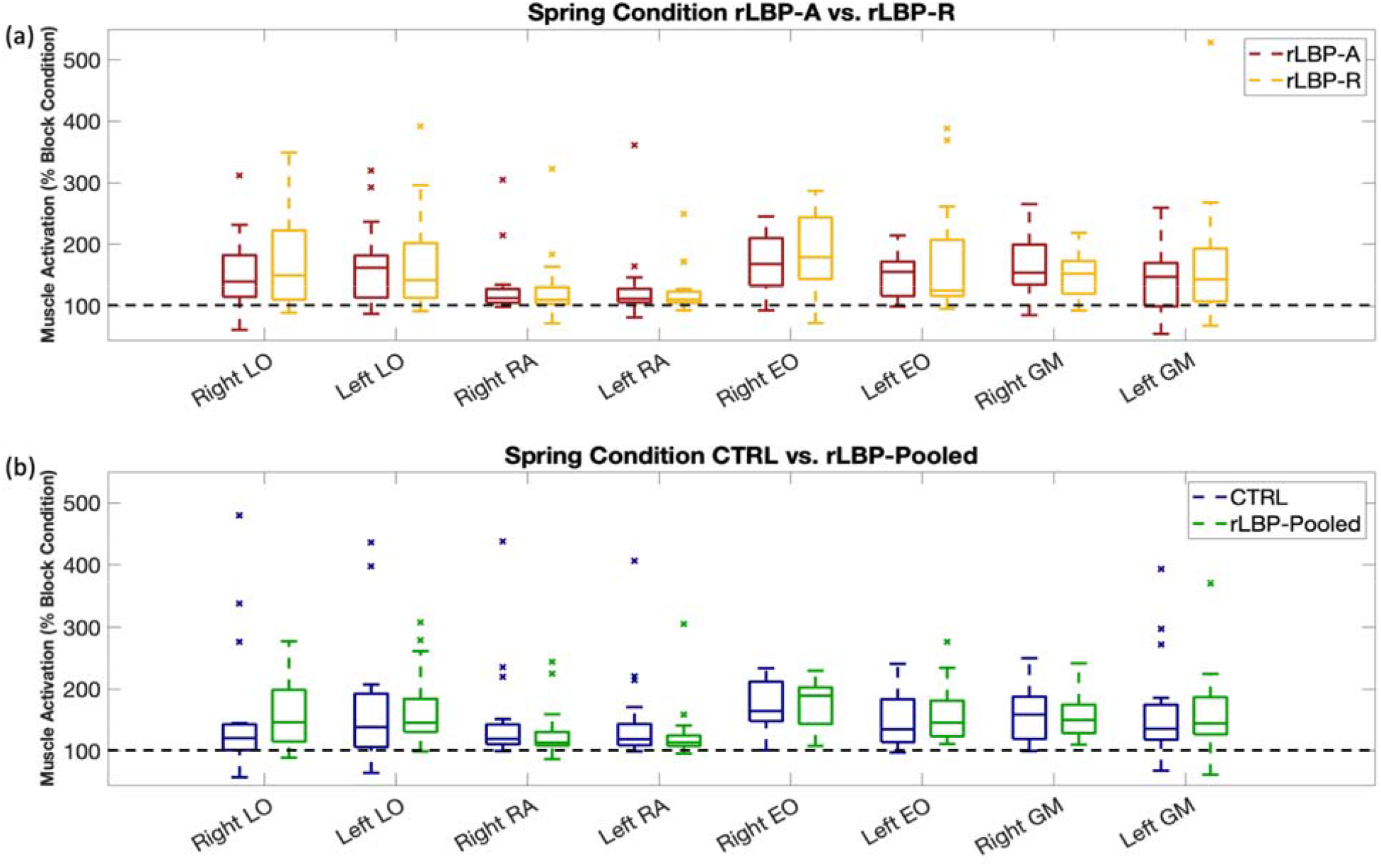
Activation amplitude during the spring condition trials across all muscles. (a) Comparison of activation amplitude between rLBP-A and rLBP-R. (b) Comparison of activation amplitude between rLBP-Pooled and CTRL. Electromyography signals were normalized to the block condition, and the black dotted lines on both graphs represent 100% muscle activation which is equivalent to the block condition trials. rLBP-A, recurrent low back pain in active pain; rLBP-R, recurrent low back pain in remission; CTRL, control; R, right; L, left; LO, Longissimus; RA, Rectus Abdominis; EO, External Oblique; GM, Gluteus Medius.

### 3.4 Muscle Co-activation

Bilateral LO, bilateral EO, bilateral GM, Right LO-Right RA, and Left LO-Left RA exhibited greater co-activation in the block condition than in the spring condition (p<.001) (Fig 6(a), Table 3). This difference was not found in Bilateral RA (Fig 6(a), Table 3).

**Table 3.** Muscle co-activation statistical results.

|  | Group Effects (p-values) |  |  | Interaction Effects (p-values) |  |  |
| --- | --- | --- | --- | --- | --- | --- |
|  | LBP | rLBP-Pooled vs. CTRL |  | LBP | rLBP-Pooled vs. CTRL |  |
| Bilateral LO | .218 | .353 |  | .279 | .459 |  |
| Bilateral RA | .380 | .260 |  | .897 | .868 |  |
| Bilateral GM | .099 | .786 |  | .960 | .132 |  |
| Bilateral EO | **<.001** | rLBP-A vs. CTRL | rLBP-R vs. CTRL | **.003** | rLBP-A vs. CTRL | rLBP-R vs. CTRL |
|  |  | .968 | **.008** |  | .606 | .070 |
| R LO – R RA | .341 | .572 |  | .427 | .830 |  |
| L LO – L RA | .347 | .699 |  | .081 | .914 |  |

Bolded font indicates $p < 0.05$

**Fig 6:**
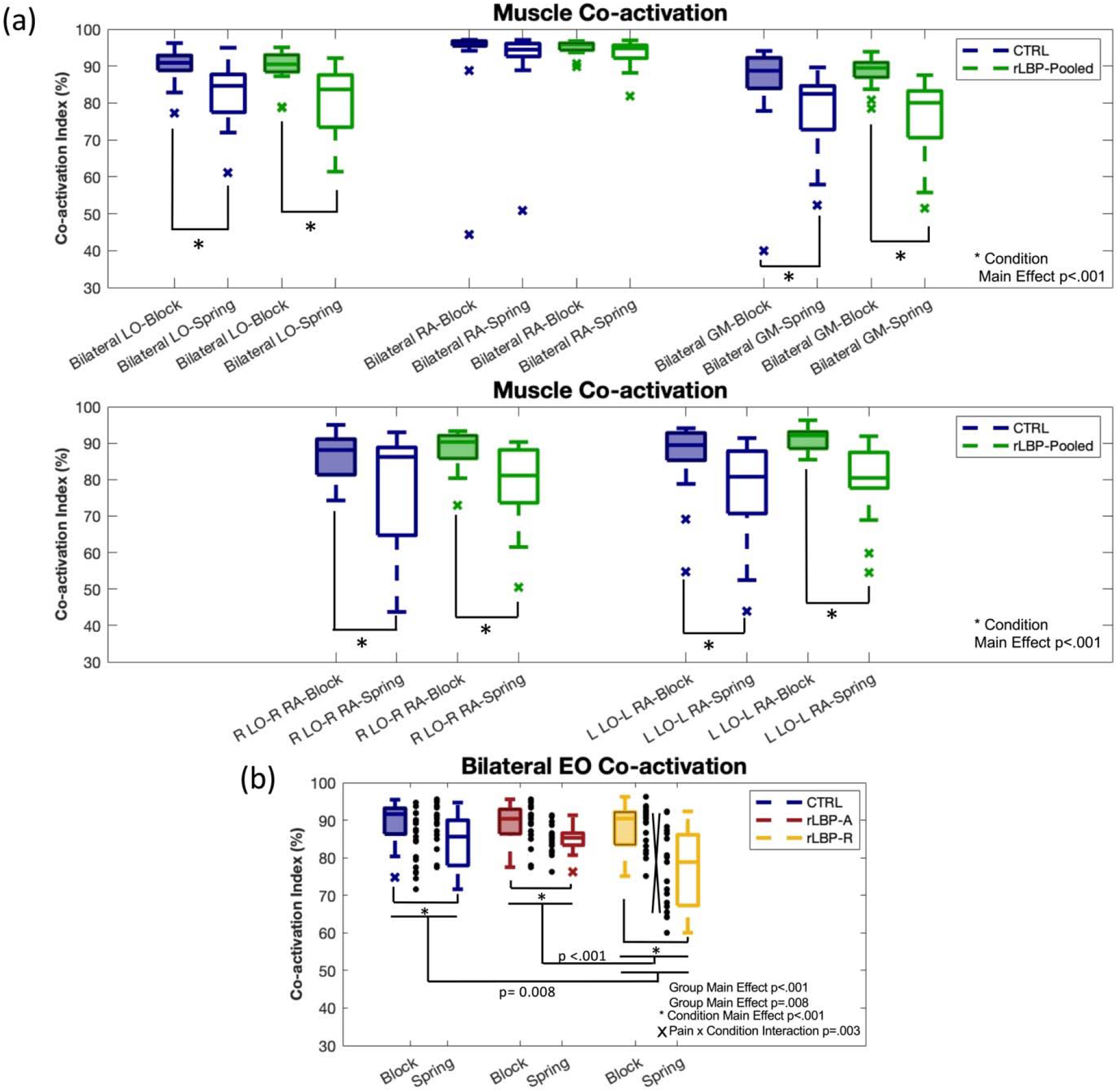
Co-activation values across examined trunk muscle pairings. (a) Co-activation values of all muscle pairings between rLBP-Pooled and CTRL, excluding bilateral EO. (b) A more detailed figure of Right RA-Right GM co-activation. (c) Co-activation value of bilateral EO. CTRL, control; rLBP-Pooled, pooled recurrent low back pain; rLBP-A, recurrent low back pain in active pain; rLBP-R, recurrent low back pain in remission; R, right; L, left; LO, Longissimus; RA, Rectus Abdominis; GM, Gluteus Medius; EO, External Oblique.

rLBP-R exhibited significantly less co-activation of bilateral EO than rLBP-A (Group main effect p<.001), as well as a greater difference in co-activation between block and spring conditions compared to rLBP-A (Interaction effect p=.003) (Fig 6(b), Table 3). There was also significantly less bilateral EO co-activation in rLBP-R than in CTRL (Group main effect p=.008), and no difference was found between CTRL and rLBP-A (Group main effect p=.968) (Fig 6(b), Table 3).

## 4. Discussion

We compared motor control, including kinematics and muscle activity, in young adults with recurrent LBP in and out of pain as well as back-healthy controls during the BDT. We found that rLBP-R had lower COP velocity on the supporting limb during the BDT compared to rLBP-A and CTRL. However, contrary to our hypothesis, there were no differences in frontal plane trunk coupling between rLBP-A, rLBP-R, and CTRL. Amplitude of trunk muscle activation was higher in the unstable spring condition compared to the stable block condition during the BDT, but there were no differences between groups. Consistent with our hypothesis, rLBP-R exhibited reduced co-activation of bilateral EO compared to rLBP-A and CTRL.

Performance of the BDT can be analyzed from the dexterity and balance perspectives. The dexterity performance reflects force control of the dexterity limb on the spring, and there were no differences between how much and how consistent participants were able to compress the spring between rLBP-A, rLBP-R, and CTRL. This result enabled us to compare other motor control variables without considering potential difference in task performance. From the balance perspective, rLBP-R exhibited a lower COP velocity compared to rLBP-A and CTRL. We first considered a practice effect, which could be present because participants were always tested first during active pain and then during remission. However, we performed a post-hoc exploratory analysis and found no evidence for a practice effect (Supplementary Material).

Without evidence of a practice effect, the lower COP velocity could be explained by a “freeze” strategy for postural control during the remission period. Jones et al. found a similar “freeze” strategy in people with LBP who were not in an active pain episode evidenced by enhanced trunk and ankle muscle activation in response to floor perturbation.^29^ It is possible that this “freeze” strategy was reflected as lower COP velocity during the BDT and could be explained by psychosocial influences. Persons with rLBP commonly experience fear avoidance and kinesiophobia,^22^ and could perceive performing BDT as a risk of inducing pain due to loss of balance. Similar “freeze” postural control was observed in height-induced postural threat.^30^ However, when individuals were in an active pain episode, pain-induced sensory noise may have resulted an increased COP velocity compared to a pain-free state. Therefore, the effect of a “freeze” strategy and pain-induced sensory noise may have cancelled out and prevented a difference in COP velocity between rLBP-A and CTRL.

Even though we observed that the rLBP group reduced trunk coupling during the BDT compared to the stable block condition while this effect was not present in the CTRL group, we did not find statistically significant differences between groups or pain status. These results differed from Rowley et al, which found that LBP-R patients exhibited reduced trunk coupling compared to the healthy control group and suggested that LBP patients may utilize a “loose” control strategy.^14^ In the current study, participants were given fewer practice trials (3 vs. 5 in Rowley et al) before the recorded trial and may have used a “tighter” control strategy while continuing to become familiar with the task. Additionally, the current study defined recurrent LBP as having less than half the days in pain in the past 6 months, whereas the Rowley study defined it as having 2 or more episodes in the past year. These discrepancies in methodological details and participant characteristics may have contributed to the different outcomes between the studies, and highlights the heterogeneity of the LBP population.

There is overall higher muscle activity during the BDT in the spring condition compared to the block condition across both groups. This is not surprising given the unstable perturbation during the spring condition, requiring greater muscular control. We also found that rLBP-R exhibited less bilateral EO co-activation compared to rLBP-A and CTRL. The bilateral EO co-activation results could indicate that when rLBP patients are in symptom remission, they adopt a “loose” strategy, while in active pain they exhibit a “tighter” control strategy, even though the co-activation was not greater than the control group. This may be an attempt to protect the spine again shearing forces since bilateral EO co-activation creates opposing forces for side-bend and rotation actions, which contributes to the frontal and transverse plane stability, respectively. The frontal and transverse planes are the least stable by design of the task,^12^ which highlights the critical role of bilateral EO and may explain the significant findings with this muscle pairing.

Further, because the calculation of co-activation is driven by both reciprocal activation timing and amplitude, and amplitude was higher in the spring condition than the block condition, the observed reduction in co-activation in the spring condition was primarily due to less synchronous timing between the two muscles. With this interpretation in mind, the greater reduction in bilateral EO co-activation during the spring condition compared to the block condition in rLBP-R compared to rLBP-A could indicate that rLBP-A exhibited a more prolonged co-activation pattern rather than rapid reciprocal patterns utilized by rLBP-R during the spring condition. Our findings highlight the important role of EO during the BDT and the influence of pain on the underlying motor control strategies adopted by patients at different time points in the course of rLBP.

There are several limitations to this study. First, we recruited minimally disabled young adults with mild to moderate pain, which may not be generalizable to other populations with LBP. Second, our definition was adapted from the National Institutes of Health Task Force definition of chronic LBP,^31^ and, therefore, may differ from other studies. Furthermore, the BDT has limited ecological validity as the task does not replicate daily activities. The BDT was chosen because it provides a continues, submaximal perturbation.^12,14^ With these limitations in mind, our findings provide insight into the different trunk control strategies utilized by individuals with rLBP. More research is needed to elucidate potential subgroups of patients who may exhibit different control strategies, and to systematically examine how different task demands influence control strategies in people with LBP.

Overall, our findings support that this cohort of individuals with rLBP demonstrates a “loose” motor control strategy in response to a continuously unstable perturbation, especially during symptom remission. The potentially increased shearing forces of the spinal segments with a “loose” control strategy may make individuals with rLBP in remission more vulnerable to another flare-up episode. Our current findings indicate that although the overall “loose” motor control demonstrated by individuals with rLBP may be “tightened” by the presence of active pain, the motor control during active pain does not appear to be “tighter” than back-healthy individuals, at least in this submaximal unstable balance task. Further investigation is needed to elucidate which aspects of trunk control in individuals with rLBP is associated with future recurrences, and its complicated interactions with task threat and pain status.

## Data Availability

All data produced in the present study are available upon reasonable request to the authors

## Acknowledgment

This study was funded by the International Society of Biomechanics Matching Dissertation Grant awarded to H-JSS. LVD was supported by grant NIH/NICHD/NCMRR R01 HD 047709. The funding body had no role in the study design, collection, analysis, or interpretation of data.

## References

1. Freburger JK, Holmes GM, Agans RP, et al. The Rising Prevalence of Chronic Low Back Pain. Arch Intern Med. 2009;169(3):251. doi:10.1001/archinternmed.2008.543

2. Hoy D, March L, Brooks P, et al. The global burden of low back pain: estimates from the Global Burden of Disease 2010 study. Ann Rheum Dis. 2014;73(6):968–974. doi:10.1136/annrheumdis-2013-204428

3. Hancock MJ, Maher CM, Petocz P, et al. Risk factors for a recurrence of low back pain. Spine J. 2015;15(11):2360–2368. doi:10.1016/j.spinee.2015.07.007

4. Hartvigsen J, Hancock MJ, Kongsted A, et al. What low back pain is and why we need to pay attention. The Lancet. 2018;391(10137):2356–2367. doi:10.1016/S0140-6736(18)30480-X

5. da Silva T, Mills K, Brown BT, Herbert RD, Maher CG, Hancock MJ. Risk of Recurrence of Low Back Pain: A Systematic Review. J Orthop Sports Phys Ther. 2017;47(5):305–313. doi:10.2519/jospt.2017.7415

6. Steffens D, Ferreira ML, Latimer J, et al. What Triggers an Episode of Acute Low Back Pain? A Case-Crossover Study: Triggers for Low Back Pain. Arthritis Care Res. 2015;67(3):403–410. doi:10.1002/acr.22533

7. van Dieën JH, Reeves NP, Kawchuk G, van Dillen LR, Hodges PW. Motor Control Changes in Low Back Pain: Divergence in Presentations and Mechanisms. J Orthop Sports Phys Ther. 2019;49(6):370–379. doi:10.2519/jospt.2019.7917

8. van Dieën JH, Kingma I, van der Bug JCE. Evidence for a role of antagonistic cocontraction in controlling trunk stiffness during lifting. J Biomech. 2003;36(12):1829–1836. doi:10.1016/S0021-9290(03)00227-6

9. Hodges P, van den Hoorn W, Dawson A, Cholewicki J. Changes in the mechanical properties of the trunk in low back pain may be associated with recurrence. J Biomech. 2009;42(1):61–66. doi:10.1016/j.jbiomech.2008.10.001

10. Marich AV, Hwang C, Sorensen CJ, van Dillen LR. Examination of the Lumbar Movement Pattern during a Clinical Test and a Functional Activity Test in People with and without Low Back Pain. PM&R. 2020;12(2):140–146. doi:10.1002/pmrj.12197

11. Freddolini M, Strike S, Lee RYW. Stiffness properties of the trunk in people with low back pain. Hum Mov Sci. 2014;36:70–79. doi:10.1016/j.humov.2014.04.010

12. Rowley KM, Gordon J, Kulig K. Characterizing the balance-dexterity task as a concurrent bipedal task to investigate trunk control during dynamic balance. J Biomech. 2018;77:211–217. doi:10.1016/j.jbiomech.2018.07.014

13. Lyle MA, Valero-Cuevas FJ, Gregor RJ, Powers CM. The lower extremity dexterity test as a measure of lower extremity dynamical capability. J Biomech. 2013;46(5):998–1002. doi:10.1016/j.jbiomech.2012.11.058

14. Rowley KM, Smith JA, Kulig K. Reduced Trunk Coupling in Persons With Recurrent Low Back Pain Is Associated With Greater Deep-to-Superficial Trunk Muscle Activation Ratios During the Balance-Dexterity Task. J Orthop Sports Phys Ther. 2019;49(12):887–898. doi:10.2519/jospt.2019.8756

15. Hodges PW, Tucker K. Moving differently in pain: A new theory to explain the adaptation to pain. Pain. 2011;152(3):S90–S98. doi:10.1016/j.pain.2010.10.020

16. Smith JA, Kulig K. Trunk–pelvis coordination during turning: A cross sectional study of young adults with and without a history of low back pain. Clin Biomech. 2016;36:58–64. doi:10.1016/j.clinbiomech.2016.05.011

17. Shih HJS, Van Dillen LR, Kutch JJ, Kulig K. Individuals with recurrent low back pain exhibit further altered frontal plane trunk control in remission than when in pain. Clin Biomech. 2021;87:105391. doi:10.1016/j.clinbiomech.2021.105391

18. Ostelo RWJG, Deyo RA, Stratford P, et al. Interpreting Change Scores for Pain and Functional Status in Low Back Pain: Towards International Consensus Regarding Minimal Important Change. Spine. 2008;33(1):90–94. doi:10.1097/BRS.0b013e31815e3a10

19. Shih HJS, Gordon J, Kulig K. Trunk control during gait: Walking with wide and narrow step widths present distinct challenges. J Biomech. 2021;114:110135. doi:10.1016/j.jbiomech.2020.110135

20. Hallal PC, Victora CG. Reliability and validity of the International Physical Activity Questionnaire (IPAQ). Med Sci Sports Exerc. 2004;36(3):556. doi:doi: 10.1249/01.mss.0000117161.66394.07

21. Fritz JM, Irrgang JJ. A comparison of a modified Oswestry Low Back Pain Disability Questionnaire and the Quebec Back Pain Disability Scale. Phys Ther. 2001;81(2):776–788. doi:10.1093/ptj/81.2.776

22. Waddell G, Newton M, Henderson I, Somerville D, Main CJ. A Fear-Avoidance Beliefs Questionnaire (FABQ) and the role of fear-avoidance beliefs in chronic low back pain and disability. Pain. 1993;52(2):157–168. doi:10.1016/0304-3959(93)90127-B

23. Winter DA, Patla AE, Frank JS. Assessment of balance control in humans. Med Prog Technol. 1990;16(1-2):31–51.

24. Schinkel-Ivy A, Nairn BC, Drake JDM. Investigation of trunk muscle co-contraction and its association with low back pain development during prolonged sitting. J Electromyogr Kinesiol. 2013;23(4):778–786. doi:10.1016/j.jelekin.2013.02.001

25. Dankaerts W, O’Sullivan PB, Burnett AF, Straker LM, Danneels LA. Reliability of EMG measurements for trunk muscles during maximal and sub-maximal voluntary isometric contractions in healthy controls and CLBP patients. J Electromyogr Kinesiol. 2004;14(3):333–342. doi:10.1016/j.jelekin.2003.07.001

26. Falconer K, Winter DA. Quantitative assessment of co-contraction at the ankle joint in walking. Electromyogr Clin Neurophysiol. 1985;25(2-3):135–149.

27. Rinaldi M, D’Anna C, Schmid M, Conforto S. Assessing the influence of SNR and pre-processing filter bandwidth on the extraction of different muscle co-activation indexes from surface EMG data. J Electromyogr Kinesiol. 2018;43:184–192. doi:10.1016/j.jelekin.2018.10.007

28. R Core Team. R: A Language and Environment for Statistical Computing. Published online 2018. https://www.R-project.org

29. Jones SL, Henry SM, Raasch CC, Hitt JR, Bunn JY. Individuals with non-specific low back pain use a trunk stiffening strategy to maintain upright posture. J Electromyogr Kinesiol. 2012;22(1):13–20. doi:10.1016/j.jelekin.2011.10.006

30. Cleworth TW, Chua R, Inglis JT, Carpenter MG. Influence of virtual height exposure on postural reactions to support surface translations. Gait Posture. 2016;47:96–102. doi:10.1016/j.gaitpost.2016.04.006

31. Deyo RA, Dworkin SF, Amtmann D, et al. Report of the NIH Task Force on Research Standards for Chronic Low Back Pain. J Pain. 2014;15(6):569–585. doi:10.1016/j.jpain.2014.03.005

