## Supplemental Material for "Trunk control in and out of an episode of recurrent low back pain during the Balance-Dexterity Task"

**Supplementary Material**

We calculated whether there is a relationship between the change in COP velocity from test to re-test (re-test minus test) and the number of days between testing to indirectly examine learning effect of the task. Note that days to retest is based on both symptom criteria on pain score as well as scheduling convenience and thus does not represent the number of days continuously in pain.

If there was a learning effect, we would expect COP velocity to be reduced from test to re-test, resulting in a negative value on the y-axis when days to retest is small, while trending towards a return to no difference in COP velocity (0 on the y-axis) when days to retest gets larger. This learning effect would then present in a trend of positive relationship in the graphs we made.

Our results did not show a significant association between the change in COP velocity from test to re-test and the number of days between testing, both in the block and spring condition. Therefore, we did not find evidence of a learning effect of COP velocity in the BDT.

### Difference in COPvel-block against Days to retest

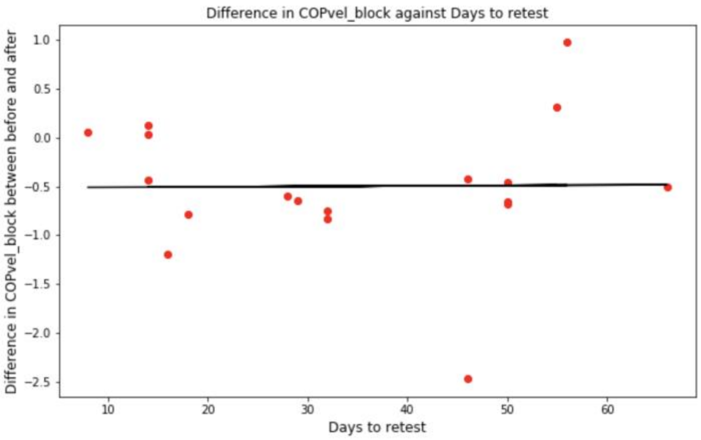


| | coef | std err | t | P>\|t\| | [0.025 | 0.975] |
|---|---|---|---|---|---|---|
| const | -0.5135 | 0.382 | -1.343 | 0.198 | -1.324 | 0.297 |
| Days to Retest | 0.0005 | 0.010 | 0.047 | 0.963 | -0.020 | 0.021 |

### Difference in COPvel-spring against Days to retest

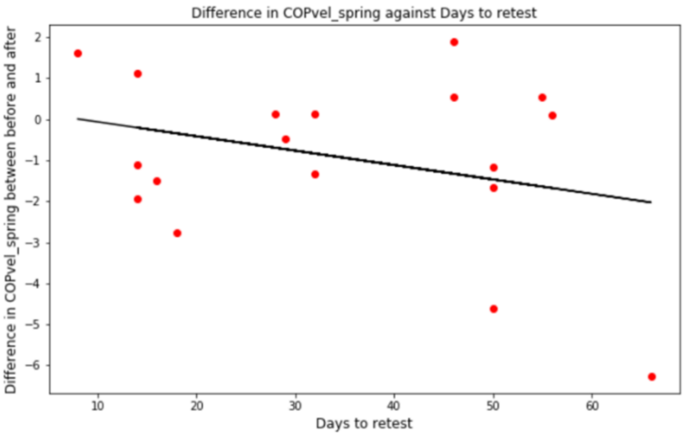


| | coef | std err | t | P>\|t\| | [0.025 | 0.975] |
|---|---|---|---|---|---|---|
| const | 0.2876 | 1.070 | 0.269 | 0.792 | -1.980 | 2.556 |
| Days to Retest | -0.0351 | 0.028 | -1.273 | 0.221 | -0.093 | 0.023 |
